# Adjusting non-pharmaceutical interventions based on hospital bed capacity using a multi-operator differential evolution

**DOI:** 10.1101/2022.07.17.22277729

**Authors:** Victoria May P. Mendoza, Renier Mendoza, Jongmin Lee, Eunok Jung

**Affiliations:** Department of Mathematics, Konkuk University, Seoul, 05029, Republic of Korea; Institute of Mathematics, University of the Philippines Diliman, Quezon City, 1101, Philippines

**Keywords:** COVID-19, social distancing, mathematical model, metaheuristic algorithm, improved multi-operator differential evolution, optimal control

## Abstract

Without vaccines and medicine, non-pharmaceutical interventions (NPIs) such as social distancing, have been the main strategy in controlling the spread of COVID-19. Strict social distancing policies may lead to heavy economic losses, while relaxed social distancing policies can threaten public health systems. We formulate an optimization problem that minimizes the stringency of NPIs during the prevaccination and vaccination phases and guarantees that cases requiring hospitalization will not exceed the number of available hospital beds. The approach utilizes an SEIQR model that separates mild from severe cases and includes a parameter µ that quantifies NPIs. Payoff constraints ensure that daily cases are decreasing at the end of the prevaccination phase and cases are minimal at the end of the vaccination phase. Using the penalty method, the constrained minimization is transformed into a non-convex, multi-modal unconstrained optimization problem, which is solved using a metaheuristic algorithm called the improved multi-operator differential evolution. We apply the framework to determine optimal social distancing strategies in the Republic of Korea given different amounts and types of antiviral drugs. The model considers variants, booster shots, and waning of immunity. The optimal µ values show that fast administration of vaccines is as important as using highly effective vaccines. The initial number of infections and daily imported cases should be kept minimum especially if the severe bed capacity is low. In Korea, a gradual easing of NPIs without exceeding the severe bed capacity is possible if there are at least seven million antiviral drugs and the effectiveness of the drug in reducing disease severity is at least 86%. Model parameters can be adapted to a specific region or country, or other infectious disease. The framework can also be used as a decision support tool in planning practical and economic policies, especially in countries with limited healthcare resources.

**Mathematics Subject Classification:** 34A55, 34H05, 90C26, 92-10

## 1. Introduction

In the early outbreaks of COVID-19, many countries have banned public gatherings, closed down schools, restaurants, land, and sea borders, and forced people to stay at home in an attempt to curb the spread of this disease [1, 2, 3]. Few countries such as the Republic of Korea and Singapore focused their control measures on intensive contact tracing and testing [2, 4]. Without vaccines and medicine, controlling the spread of COVID-19 relied mainly on non-pharmaceutical interventions (NPIs). These control measures are essential to public health but have caused an immense burden on the social and economic aspects of life [5, 6].

During the early stages of COVID-19, most countries have monitored incidence cases and deaths and relied on this information in crafting policies [2, 3]. The development of vaccines and oral antiviral drugs greatly impacted public health by reducing the severity of infections [7]. The fast rollout of vaccines, use of highly effective vaccines and medicine, and implementation of strict social distancing policies are expected to greatly minimize infections [8, 9, 10]. However, the protection given by pharmaceutical interventions alone did not guarantee the suppression of COVID-19 as variants of the virus continued to emerge. During the highly-transmissible omicron wave, the focus of control shifted to managing severe cases and minimizing deaths. The timing of lifting and intensity of easing strict NPIs have been important policy questions during the course of the COVID-19 pandemic. To this end, mathematical models that incorporate various aspects of COVID-19 and NPIs are proving to be important decision support tools [11, 12, 13, 14].

Motivated by the events of the COVID-19 pandemic, we aim to provide a framework for planning social distancing policies based on the number of infections requiring hospitalization. The approach utilizes a Susceptible-Exposed-Infected-Isolated-Recovered (SEIQR) model that distinguishes mild from severe cases and includes a parameter that quantifies NPIs. The policy period is divided into two phases: a prevaccination phase, where only NPIs are implemented, followed by a vaccination phase, where vaccines are assumed to be available as additional control measures. We formulate optimization problems that minimize the stringency of NPIs during the prevaccination and vaccination phases and guarantee that cases requiring hospitalization will not exceed the number of available hospital beds. A measure of cost-effectiveness is presented as a basis for the choice of the frequency of policy change. To solve the optimization problems, we transform the constrained problems into unconstrained ones and implement a metaheuristic algorithm called the improved multi-operator differential evolution (IMODE) [15].

The presented framework can address key policy issues on the timing and level of adjusting NPIs. It is most relevant during the early epidemic phase, wherein pharmaceutical interventions are not yet available but managing the number of infections is a top priority. The timing of easing of NPIs is crucial at the start of vaccination in order not to compromise the benefits of vaccines [16, 17, 18]. Since a time-dependent epidemiological model is embedded in the method, data can be readily updated to fit the country or region where the framework is applied. As an application, we implement the method to forecast the optimal intensity and timing of social distancing policies in Korea. An extended, more elaborate epidemiological model is adopted that includes variants, vaccination with primary and booster shots, waning of immunity, and administration of oral antiviral drugs [19]. Optimal solutions during the vaccination phase are determined under different scenarios.

## 2. Methods

In this section, we present the epidemiological models utilized in the optimization problems. We formulate the optimization framework and transform the constrained into an unconstrained problem via the penalty method. We also present a measure of the cost-effectiveness of strategies and discuss in more detail the metaheuristic algorithm IMODE.

### 2.1. Epidemiological model

The mathematical model, illustrated in Figure 1, follows an SEIQRD structure. The subscript *v* denotes vaccination. A susceptible individual (*S*) is effectively (*V*) or ineffectively (*U*) vaccinated at a rate *θ* with vaccine effectiveness e. After an average of 1/ω days, individuals in *V* develop full immunity to the disease (*P*). Without full immunity, susceptible individuals (*S, U, V*) can become exposed (E) with a force of infection λ(*t*). An exposed individual becomes infectious (*I*) after 1/κ days on average, and is eventually confirmed and isolated in 1/α days. A confirmed individual can be classified as mild (*Q*^m^) or severe (*Q*^s^). The parameter p represents the severe rate or the proportion of infected individuals that becomes severe. Isolated individuals can recover (*R*) at rate γ^m^ or γ^s^. A proportion f of severe cases is assumed to die (*D*). Finally, we suppose that infected individuals who were vaccinated have a reduced chance of having severe symptoms by a factor of 1 *− e*^*s*^. The parameter ξ depicted by the red dashed arrow going into *E* represents the number of daily imported cases.

**Figure 1:**
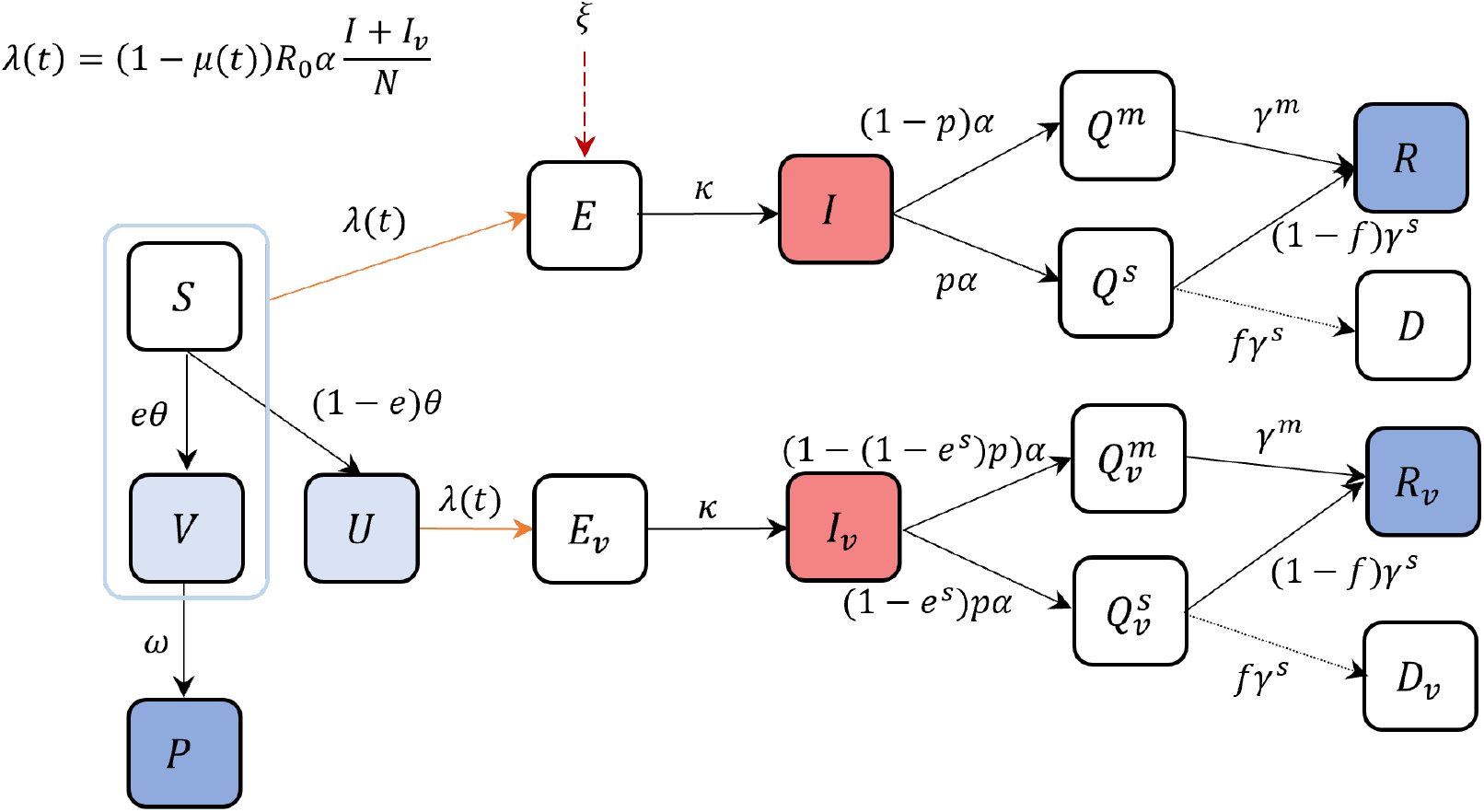
Diagram for the epidemiological model used in the optimization problem. The model follows an SEIQRD structure and considers vaccination, denoted by the subscript *v*. The compartments *V, U*, and *P* represent the effectively vaccinated, ineffectively vaccinated, and fully protected groups, respectively, while the isolated mild and severe groups are denoted *Q*^m^ and *Q*^s^, respectively. The other subclasses are susceptible (*S*), exposed (*E*), infectious (*I*), recovered (*R*), and death (*D*).

The susceptible and vaccinated compartments are described by the following differential equations

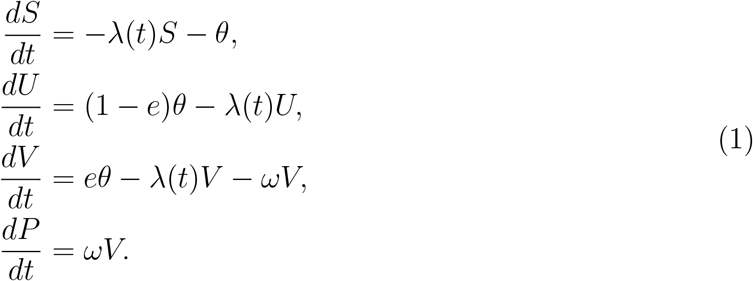

The infected, recovered, and death compartments are given by

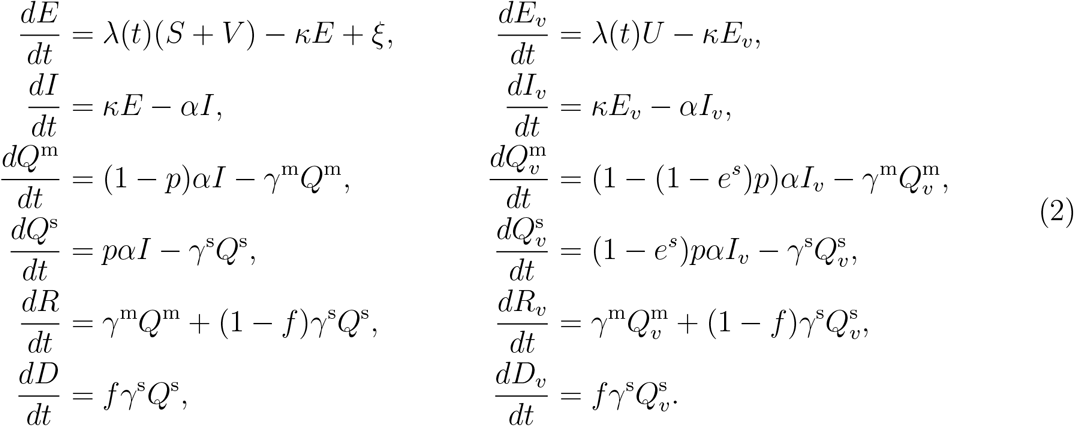

The force of infection λ(*t*) is defined as

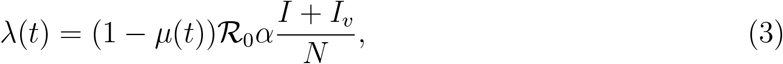

where

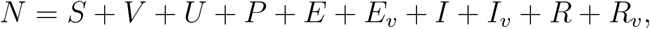

*µ*(*t*) is a time-dependent parameter representing the intensity of NPIs, and ℛ_0_ is the basic reproductive number. The parameter *µ*(*t*) takes a value between 0 and 1. A *µ* value close to 1 translates to a high reduction in transmission, which is assumed to result from a strict implementation of NPIs. On the other hand, a *µ* value close to 0 implies more relaxed NPIs. In the prevaccination phase, we simply set the speed of vaccination θ to zero and the model reduces to a single SEIQRD flow. Because vaccines have different effectiveness, we considered low and high values for e and *e*^*s*^. If the number of daily imported cases ξ is set to 0, it means that screening measures at the border detect all non-locally transmitted cases. To investigate the effect of importation on the spread of the disease, we also perform simulations on varying ξ. Description of the model parameters and their values are listed in Table 1. The initial susceptible, exposed E_0_, and infectious *I*_0_ population are set to 999945, 50, and 5, respectively. The rest of the state variables are initially set to zero. The initial total population N_0_ is 1000000.

**Table 1:** Description and values of the parameters in the epidemiological model.

| Parameter | Description (unit) | Value | Ref |
| --- | --- | --- | --- |
| $\mathcal{R}_0$ | Basic reproductive number | 3.17 | [20, 21, 22] |
| $1/\alpha$ | Mean infectious period (day) | 4 | [23, 24] |
| $1/\omega$ | Mean period to develop full immunity (day) | 40 | Calculated |
| $1/\kappa$ | Mean latent period (day) | 4 | [23, 25] |
| $p$ | Proportion of infections that becomes severe | 2.28% | [26] |
| $1/\gamma^m$ | Mean duration of hospitalization for mild cases (day) | 11.7 | [27] |
| $1/\gamma^s$ | Mean duration of hospitalization for severe cases (day) | 14 | Assumed |
| $f$ | Mean fatality rate among severe cases | 0.439 | Calculated |
| $e$ | Vaccine effectiveness against infection | 0.91 (high) | [28] |
|  |  | 0.468 (low) | [29] |
| $e^s$ | Vaccine effectiveness against severe disease | 0.96 (high) | [30] |
|  |  | 0.555 (low) | [29] |
| $\xi$ | Number of imported cases | 0 | Assumed |

### 2.2. Formulation of the optimization problem

The stringency of NPIs is incorporated into the model as a factor appearing in (3) that reduces transmission by 1 *− µ*(*t*). In the optimization problem, we aim to determine the least value of *µ*(*t*) such that the sum of the severe patients 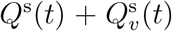 (*t*) throughout the policy period does not exceed the maximum severe bed capacity *H*_*max*_. Payoff constraints to ensure that daily cases are decreasing at the end of the prevaccination period and there are minimal cases at the end of the vaccination period are incorporated. A gradual easing of policies during the vaccination phase is also added as a constraint. The length of the prevaccination and vaccination periods and frequency of policy change are decided by the user.

Suppose that the prevaccination phase is divided into n_1_ equal periods 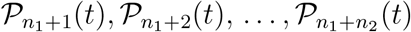 and the vaccination phase into n_2_ equal periods 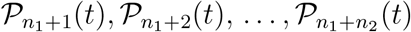. We assume that *µ*(*t*) is a piecewise constant function over disjoint periods given by

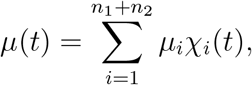

where χ_*i*_(*t*) is the characteristic function of 𝒫_*i*_(*t*), *i* = 1, 2, …, *n*_1_ + *n*_2_. Note that each *µ*_*i*_ assumes a constant value between 0.05 and 0.95. The goal of the optimization problem in the prevaccination phase is to

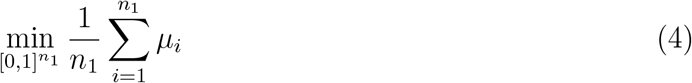

such that

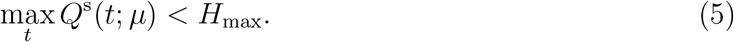

Here, *Q*^s^(*t*; *µ*) is solved from (1)-(3). The objective function in (4) simply means that we want to find the set of *µ*_*i*_ (*i* = 1, …, *n*_1_) that gives the minimum average value of all the *µ*_*i*_. The constraint in (5) ensures that the number of severe cases at any time during the prevaccination period does not exceed the threshold value *H*_*max*_ for the number of severe beds that is set by the user. Furthermore, we include a payoff constraint which guarantees that the number of cases at the end of the prevaccination phase is decreasing. That is,

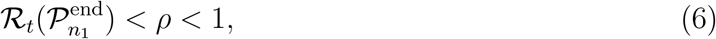

where 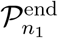 is the last time point on the period 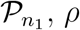 is a constant less than 1, and ℛ_*t*_(*t*) is the effective reproduction number given by

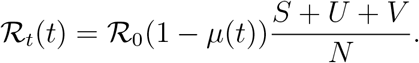

Hence, the full optimization problem for the prevaccination phase consists of (4)-(6) with the epidemiological model (1)-(3).

For the vaccination phase, we consider the following objective function formulated similarly as above,

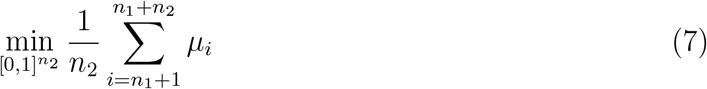

such that (5) and

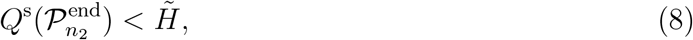

hold, where 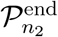 denotes the last time point on period 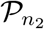. The payoff constraint (8) ensures that the number of severe patients by the end of the vaccination phase is less than a constant 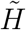, with 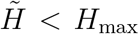. To have a gradual easing of policies during the vaccination phase, we further impose that

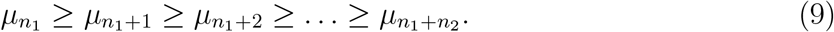

This means that successive values of *µ*_*i*_ in the vaccination phase are decreasing. The full optimization problem for the vaccination phase is given by (7)-(9), (5), and the model (1)-(3).

To solve the constrained optimization problems, we use a penalty method and convert them into unconstrained problems. Equivalently, for the prevaccination phase, we minimize over 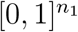 the following objective function

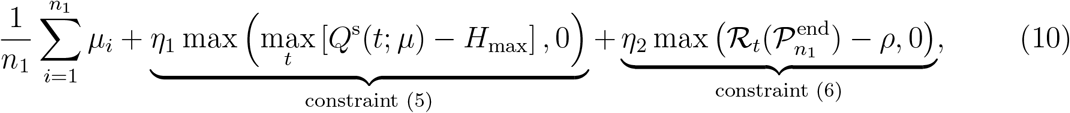

where the penalty terms η_1_, η_2_ >> 1. For the vaccination phase, the unconstrained problem is to minimize

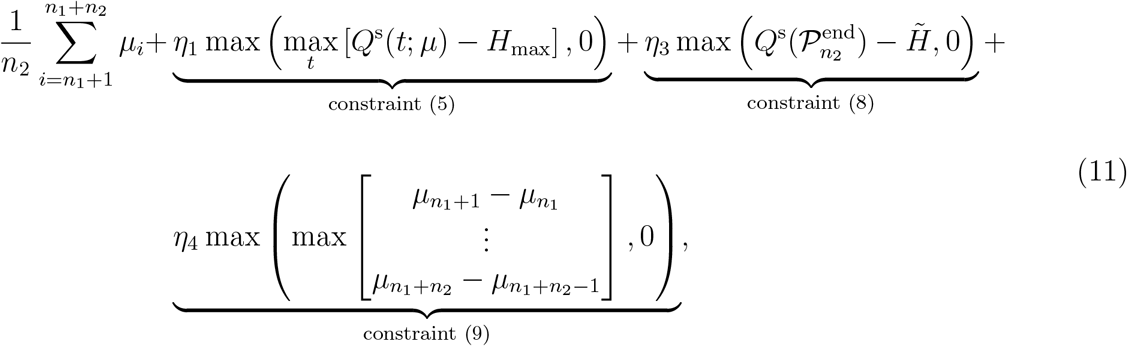

Over 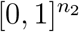, where η_3_, η_4_ >> 1. Note that the penalty terms chosen are exact [31], which means that the solution of the unconstrained problem is the same as the constrained one.

In the simulations, we assume that the entire policy period is 18 months (540 days), consisting of a 9-month prevaccination phase and a 9-month vaccination phase. In the prevaccination phase, we consider varying the frequency of policy change, that is, we set *n*_1_ = 1 (uniform), *n*_1_ = 3 (quarterly), *n*_1_ = 9 (monthly), or *n*_1_ = 18 (biweekly), or *n*_1_ = 36 (weekly). We assume that the severe bed capacity *H*_*max*_ is 100, which is 0.01% of the initial total population N_0_, and the number of severe cases by the end of the vaccination phase 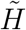 is 1% of *H*_*max*_. Moreover, vaccination is assumed to proceed at a constant rate of *θ* = 0.8N_0_/30*σ*, where *σ* is the number of months it takes to vaccinate 80% of N_0_. We consider different speeds of vaccination, that is, *σ* = 6, 9, 12, or 24 months. Because the constraints have different magnitudes relative to the value of the objective function (4), we chose appropriate weight constants *η*_*i*_. The parameter settings used in the simulations are summarized in Table 2. To solve (10) and (11), we implement the metaheuristic algorithm IMODE. Because IMODE is probabilistic, we run it 20 times and choose the solution with the least cost function value.

**Table 2:**
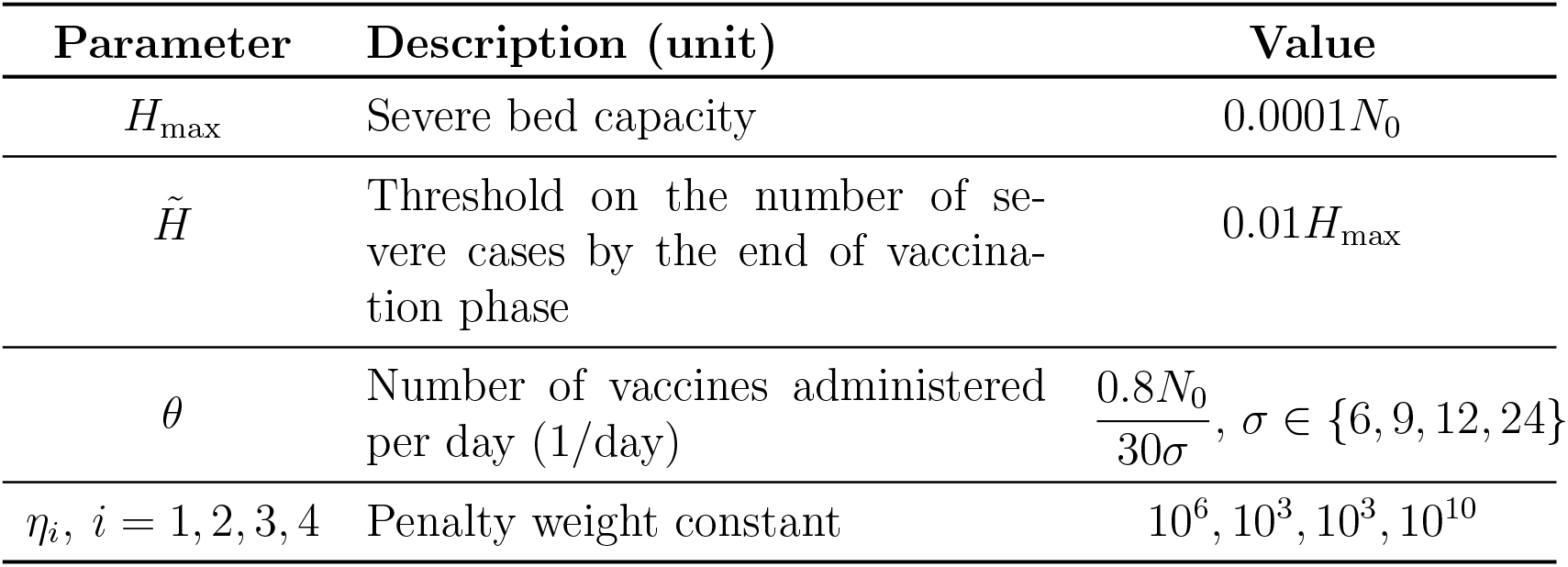
Description and values of the parameters in the optimization problem. The parameter *N*_0_ denotes the initial total population which is set to 1000000.

### 2.3. Mathematical Model of COVID-19 in Korea

As variants, booster vaccines, antiviral drugs, and waning of immunity have emerged as critical drivers of infection, a model that captures these factors is considered. We adopt the model in [19], which is an extended version of the previously presented epidemiological model to describe COVID-19 transmission in Korea.

The model illustrated in Figure 2 is divided into two parts: infection flow (top) and vaccination flow (bottom). In the vaccination flow, *S* denotes the susceptible group with no vaccine- or infection-induced immunity. Once a susceptible gets vaccinated assuming primary vaccine effectiveness *e*_1_, the individual moves to the ineffectively vaccinated *U* or effectively vaccinated *V* group. The number of people vaccinated per day is described by the parameter *θ*^*υ*^. After an average of 1/ω days, the effectively vaccinated individual moves to the protected group for the pre-delta and delta variants *P*_2_, or against all variants *P*_3_. Note that in the flow from *V* to *P*_2_ or *P*_3_, we consider a conditional probability 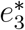, depending on the vaccine effectiveness against the different variants. That is, 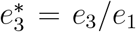. Moreover, we assume that vaccine-induced immunity of the protected groups wanes after 1/τ_*υ*_ days on average. The individuals eventually move to the waned group *W*, and then to *V*_*b*_ after getting a booster shot three or four months after completing the primary vaccines. After an average of 1/ω_*b*_ days, those who received booster shots move back to the protected groups depending on the probabilities 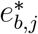 which are computed similarly as before.

**Figure 2:**
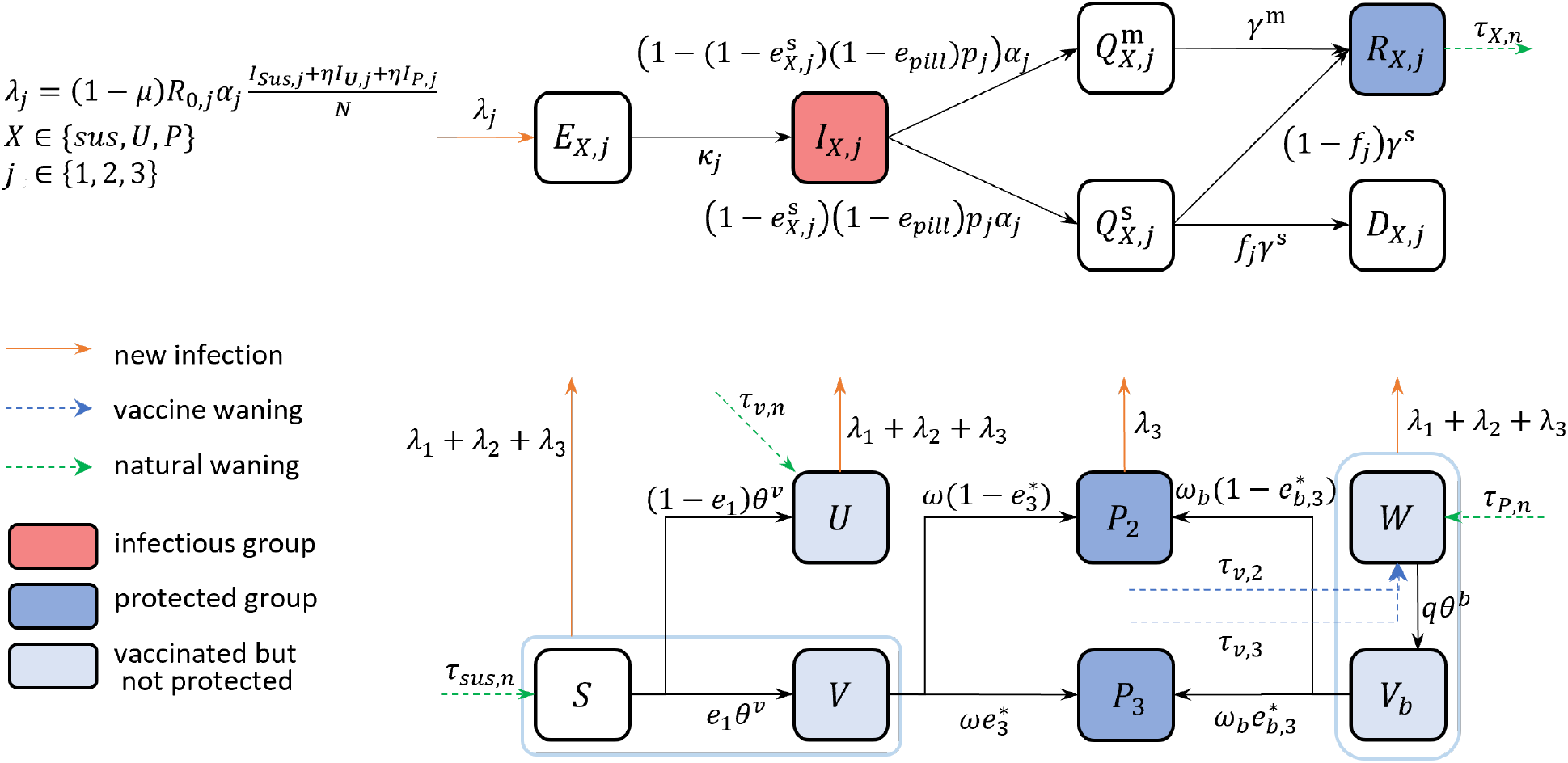
Flowchart of the COVID-19 model considering variants, waning of immunity, booster vaccines, and antiviral drugs.

Exposure to the virus occurs when a susceptible from class *X* comes in contact with an infectious individual in *I* with a force of infection λ_*j*_. The variable *X* denotes the class where the infected was from and it can be sus (from *S* or *V*), *U* (from U), or *P* (from *P*_1_, *W*, or *V*_*b*_). The subscript j denotes the variant and it can be 1 (infected by pre-delta), 2 (infected by delta), or 3 (infected by omicron). The reason for dividing the infectious group is to distinguish the different characteristics of infection, e.g. effect of vaccines for those who got infected, and the transmissibility and latent period of the variants. The *E, I, Q*^m^, *Q*^s^, *R*, and *D* classes are the same as in a previous subsection, with each class having nine variations depending on the subscripts. Furthermore, we include the impact of antiviral drugs on disease severity and the waning of immunity obtained from a previous infection. The probability of getting severe symptoms is assumed to be reduced by a factor 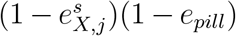. To include the waning of natural immunity depicted by the green arrow in Figure 2, individuals in *R* are assumed to have immunity only for 1/τ_*X,n*_ days on average. Note that the natural waning rate and vaccine effectiveness against severe disease are different for sus, *V*, and *P*. The system of differential equations describing the model and the list of parameters are given in the Appendix.

Before applying the optimization framework using model (A.1), we first establish the baseline value for the intensity of NPIs (*µ*) in Korea by fitting the model to the cumulative confirmed cases data from February 26, 2021, when vaccination began, until February 3, 2022, when the testing method was changed. We then forecast the number of infections given a fixed *µ* value and different amounts of antiviral drugs and compare the results to the forecasts using the optimal *µ* values during the vaccination phase, utilizing model (A.1).

The estimation period covered four distinct social distancing phases in Korea: Social Distancing level 2 (SD2) from February 26 until July 11, 2021, Social Distancing level 4 (SD4) from July 12 until October 31, 2021, Gradual Recovery (GR) from November 1 until December 17, 2021, and Suspended Gradual Recovery (SGR) from December 18, 2021 to February 3, 2022. However, these phases do not directly reflect the stringency of NPIs since there are slight differences in each social distancing phase. For example, the restriction on the number of people in a private gathering, which is an important policy in controlling transmission, changed frequently. Therefore, we divide the estimation period every two weeks and determine *µ* on each interval by fitting the model to the cumulative confirmed data using IMODE. We look for the best-fitted µ value by minimizing the following objective functional

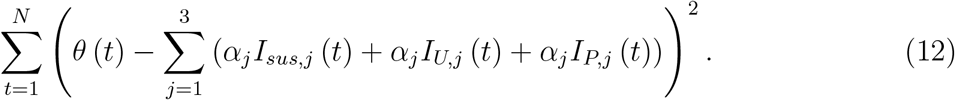

As of April 29, 2022, Korea had used 2504630 drugs (Paxlovid: 2328610, Lagevrio: 176020) and has a remaining supply of 5775670 drugs (Paxlovid: 4942710, Lagevrio: 832960) [32]. This means that the proportion of Paxlovid among the used antiviral drugs is 93%, among the reserve antiviral drugs is 86%, and the total supply is 88%. We investigate future scenarios by first considering a fixed *µ* = 0.1, 0.3, 0.5, 0.7 and setting the number of antiviral drugs to five or seven million. Then we apply the optimization framework to determine the optimal *µ* with varying amounts of antiviral drugs (five million, six million, and seven million) and proportions of Paxlovid *ϕ* (80%, 84%, 88%, 92%, 96%, and 100%) among the antiviral drugs used. We assume that the reduction in severity by Paxlovid and Lagevrio are 89% [33] and 30% [34], respectively. Simulation for the forecast starts on February 3, 2022, which is the final time of estimation, and ends on December 31, 2022. The threshold value on the number of severe cases at the end of the simulation period is set to 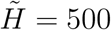.

### 2.4. Measure of cost-effectiveness

To determine the frequency of policy change, we adopt a measure of cost-effectiveness based on the cost of implementation of NPIs and the number of cases averted by the intervention strategy [35]. The cost-effectiveness ratio (CER) is used as a basis for choosing *n*_1_, *n*_2_ = 9 (monthly policy change). We also use this measure in the application of the method using COVID-19 data of Korea to compare the optimal strategies under different amounts of antiviral drugs.

The total cost of implementation of NPIs is given by 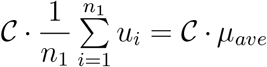, where *𝒞* is the average cost of implementation of NPIs for the whole population for *n*_1_ periods and *µ*_*ave*_ is the average of all the *µ*_*i*_ in the prevaccination phase. The CER is computed as

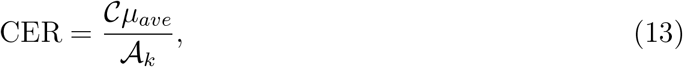

where k = 1 or 2,

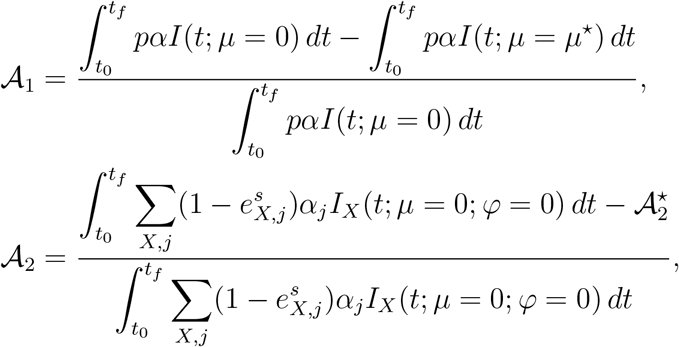

and

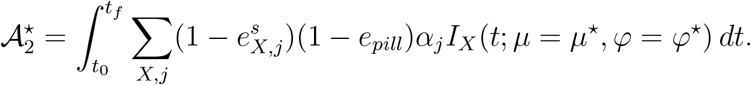

The number of averted severe cases 𝒜_*k*_ is computed as the relative difference between the number of severe cases when there are no NPIs and when there are NPIs whose intensity is quantified by *µ*^*⋆*^. In 𝒜_2_, *φ* denotes the number of antiviral drugs and *φ*^*⋆*^ can be five, six, or seven million. We use 𝒜_2_ to calculate the CER and compare the optimal strategies in Korea depending on the supply of antiviral drugs (Figure (2)). Meanwhile, 𝒜_1_ is used to calculate the CER and compare the strategies resulting from different frequencies of policy change (Figure 1). For simplicity of calculations, we set *𝒞* = 1 since it only appears as a factor in the CER in (13) and will not affect the ranking of the different policies.

### 2.5. Improved multi-operator differential evolution

In Figure 3, we observe a non-convex and multi-modal surface plot of the objective function (10) with *n*_1_ = 2. Since the surface has many local minima near the global minimum, a local optimizer is not suitable for this problem. Evolutionary algorithms have become popular because of their capability of obtaining global minimum and ease of use. They do not require the derivative of the function and will only use function evaluations. Applications of these algorithms have been explored in many areas of science and engineering [36, 37, 38, 39, 40].

**Figure 3:**
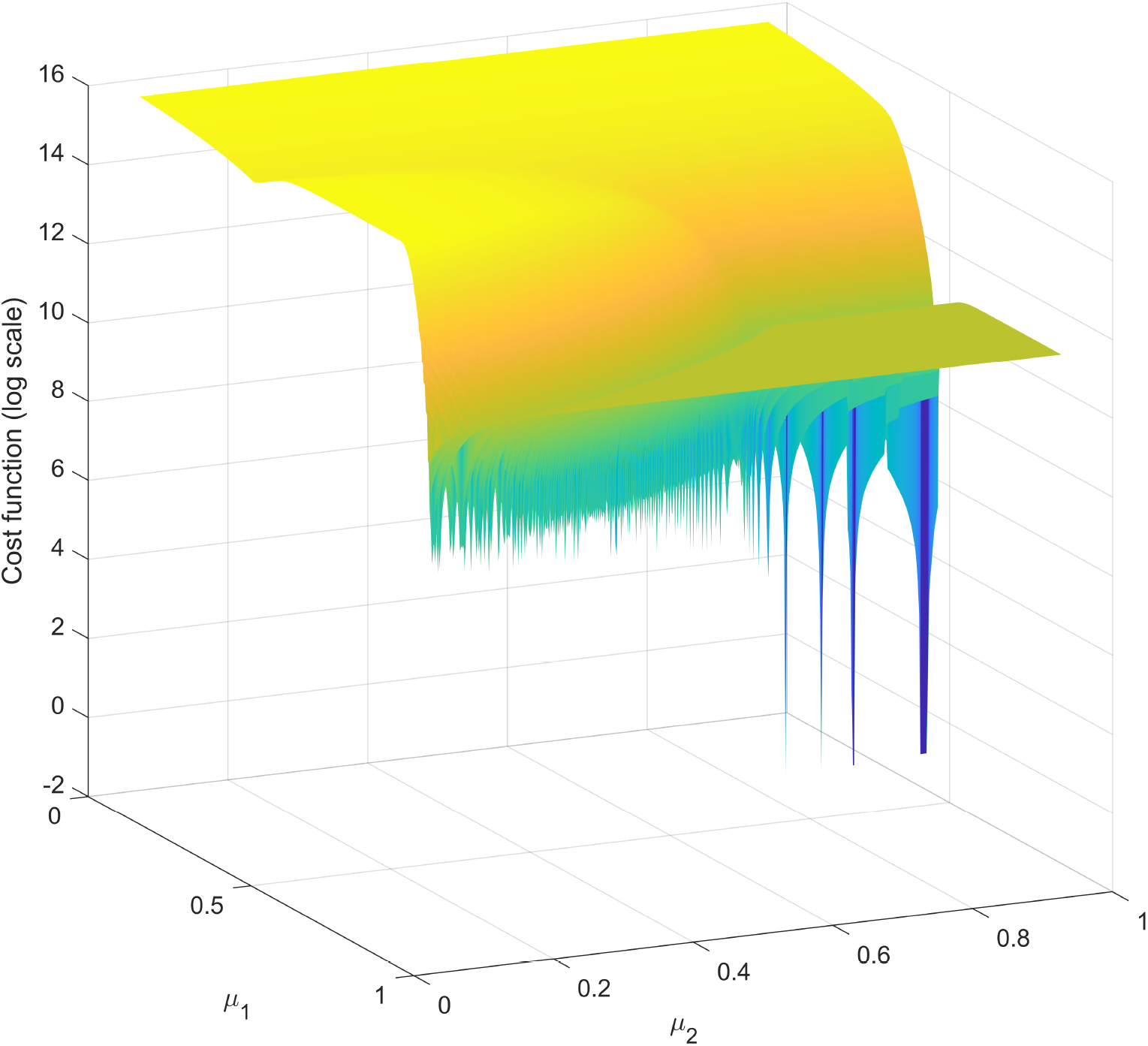
Surface plot in log-scale of the objective function in (10) for *n*_1_ = 2. The objective function is non-convex, multi-modal and has many local minima near the global minimum.

Several algorithms are continuously being developed that can obtain the global minimum with high accuracy and low computational time [41, 42, 43]. Yearly competitions are held to determine algorithms that can solve optimization problems given certain criteria and benchmark functions. In 2020, the Competition on Single Objective Bound Constrained Numerical Optimization was held and the Improved multi-operator differential evolution (IMODE) [15] ranked first during this competition. For this reason, we use IMODE to solve the minimization problems (10) and (11). Since evolutionary algorithms have been shown to be effective in estimating parameters of biological systems [44, 45, 46, 47], we also use IMODE to estimate the parameters for Korea in the COVID-19 model (A.1) by minimizing (12).

The inputs of IMODE are the objective function, the dimension of the problem, and the bound constraints. At the beginning of the IMODE process, an initial population from the search space is generated. And then, the objective function of each population member is calculated and sorted in ascending order. IMODE divides the population into several subpopulations, which are all evolved using three different mutation operators. To preserve population diversity, archiving is done. The three operators are given as follows:

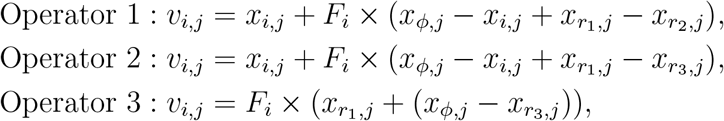

where *r*_1_, *r*_2_, *r*_3_ *≠ i* are randomly generated integers, 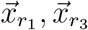 are randomly chosen from the population, 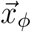 is selected from the top 10% members of the population, and 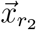 is randomly selected from the union of the archive and the population. The scaling factor *F*_*i*_ is assigned based on success-historical memory [48]. Operators 1 and 2 move the member in the current population to the best points with and without archiving, respectively [49]. Operator 3 is a weighted random-member-to-best operator [15]. The size of each sub-population (*NP*_*op*_, *op* = 1, 2, 3) is iteratively adjusted based on the diversity of the sub-populations and the quality of the solutions. The size of the population per generation (NP_*G*_) is also reduced linearly [48]. After the mutation operators are carried out, crossover are implemented randomly to create a new set of solutions. To speed up the convergence of IMODE, a local search is implemented when the number of function evaluations exceeds 85% of the maximum function evaluations (*MAX*_*FES*_). Figure 4 shows the flowchart of the IMODE algorithm. In our simulations, we used the default parameters from the original IMODE except for *MAX*_*FES*_, which we set to 20000 times the dimension of the problem. For a detailed discussion of the IMODE, we refer the readers to [15].

**Figure 4:**
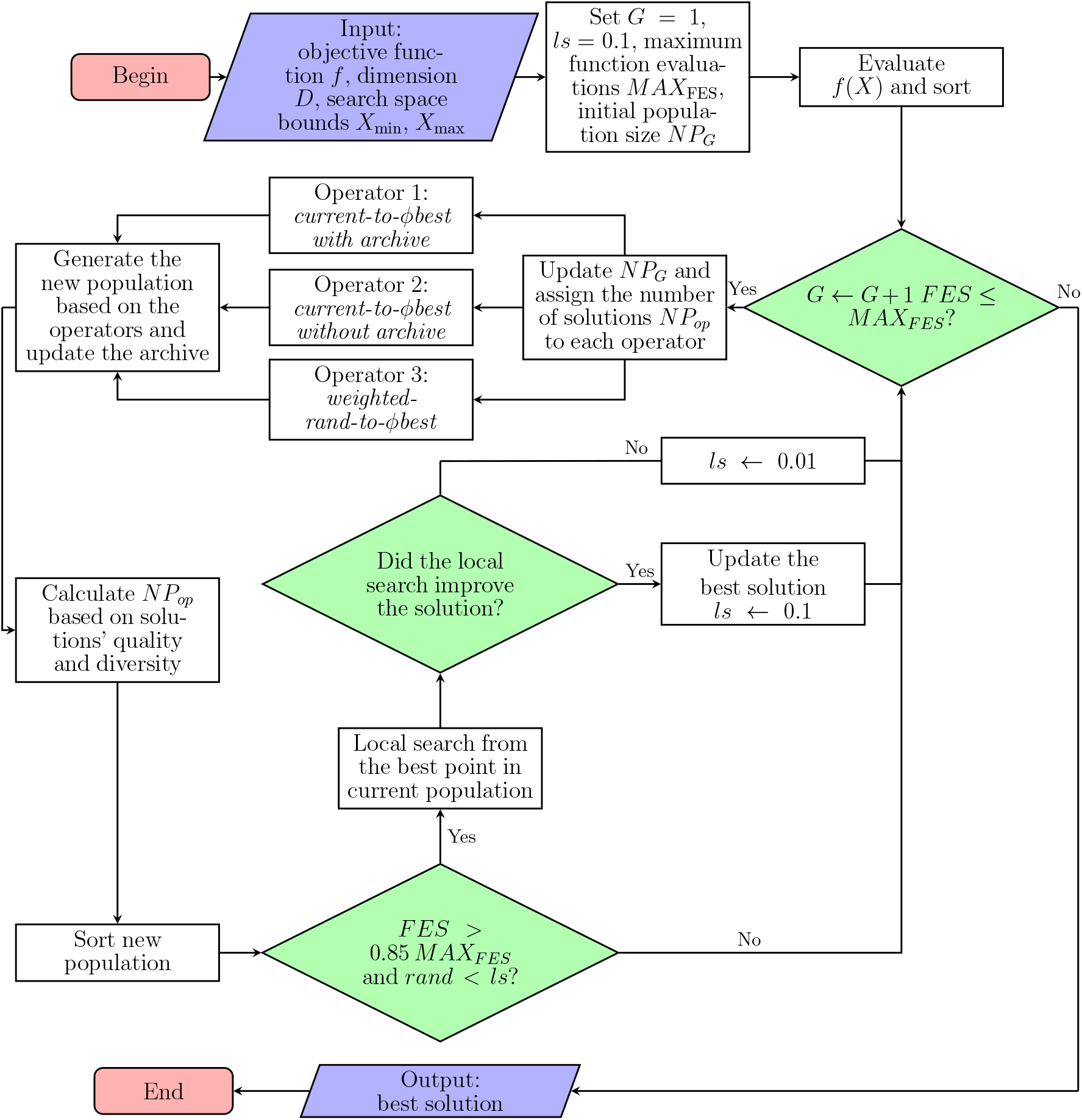
Flowchart of IMODE algorithm

## 3. Results

### 3.1. Optimal solutions and cost-effectiveness analysis during the prevaccination phase

Figure 5 shows the results of the optimization problem (10) during the prevaccination phase for different frequencies of policy change. Here we considered uniform (*n*_1_ = 1), quarterly (*n*_1_ = 3), monthly (*n*_1_ = 9), biweekly (*n*_1_ = 18), or weekly (*n*_1_ = 36) policy changes in a span of nine months. The black curves are the plots of the severe patients *Q*^s^(*t*) and the colored lines are the optimal values of µ per period.

**Figure 5:**
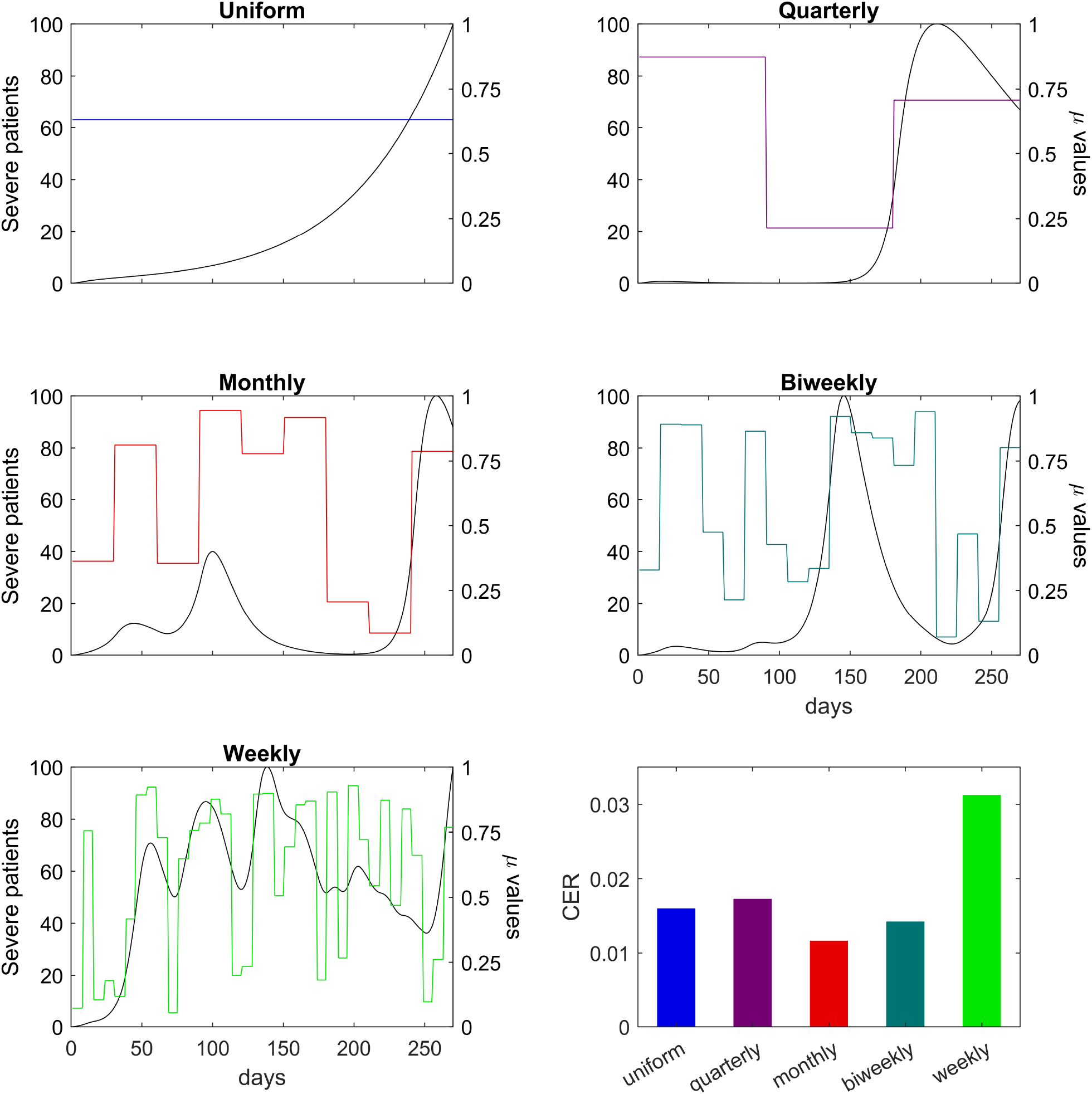
Severe patients, optimal *µ* values, and cost-effectiveness ratio (CER) for different frequencies of policy change. The policy changes once (uniform) if *n*_1_ = 1, quarterly if *n*_1_ = 3, monthly if *n*_1_ = 9, biweekly if *n*_1_ = 18, or weekly if *n*_1_ = 36. The black curves are the severe patients *Q*^s^(*t*), while the colored lines correspond to the optimal values of *µ*. The bottom right panel shows the CER for the different policies.

In the uniform policy, the optimal value of µ that should be maintained throughout the prevaccination phase is 0.630. The number of severe patients grew exponentially, almost reaching *H*_*max*_ = 100 by the end of this phase. Having two constraints on a one-dimensional optimization problem may result in an infeasible solution. For this reason, although the severe patients did not exceed *H*_*max*_, the payoff constraint, which ensures that incidence cases are decreasing by the end of the period, is not satisfied.

If the policy is changed every three months, the resulting optimal values of µ on the three consecutive periods are 0.873, 0.214, and 0.708. If the policy is changed monthly, the optimal values of µ on the consecutive months are 0.363, 0.812, 0.355, 0.944, 0.778, 0.917, 0.207, 0.085, and 0.787. The optimal solutions for the biweekly and weekly policies are illustrated in Figure 5. The average values of µ and the corresponding CER for the different policies are calculated using (13). The resulting CER are 0.016, 0.017, 0.012, 0.014, and 0.031 and are shown at the bottom right panel in Figure 5. In the succeeding simulations, we set n_1_, n_2_ = 9, which translates to a monthly policy change.

### 3.2. Optimal policy strategies during the vaccination phase

Figure 6 shows the optimal solutions considering high or low vaccine effectiveness (*e* and *e*^*s*^) and speeds of vaccination depending on the value of σ. The plots of the number of severe patients 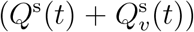 are also shown. The solid curves denote the results using a highly effective vaccine, while the dashed curves denote the results using a low effective vaccine.

**Figure 6:**
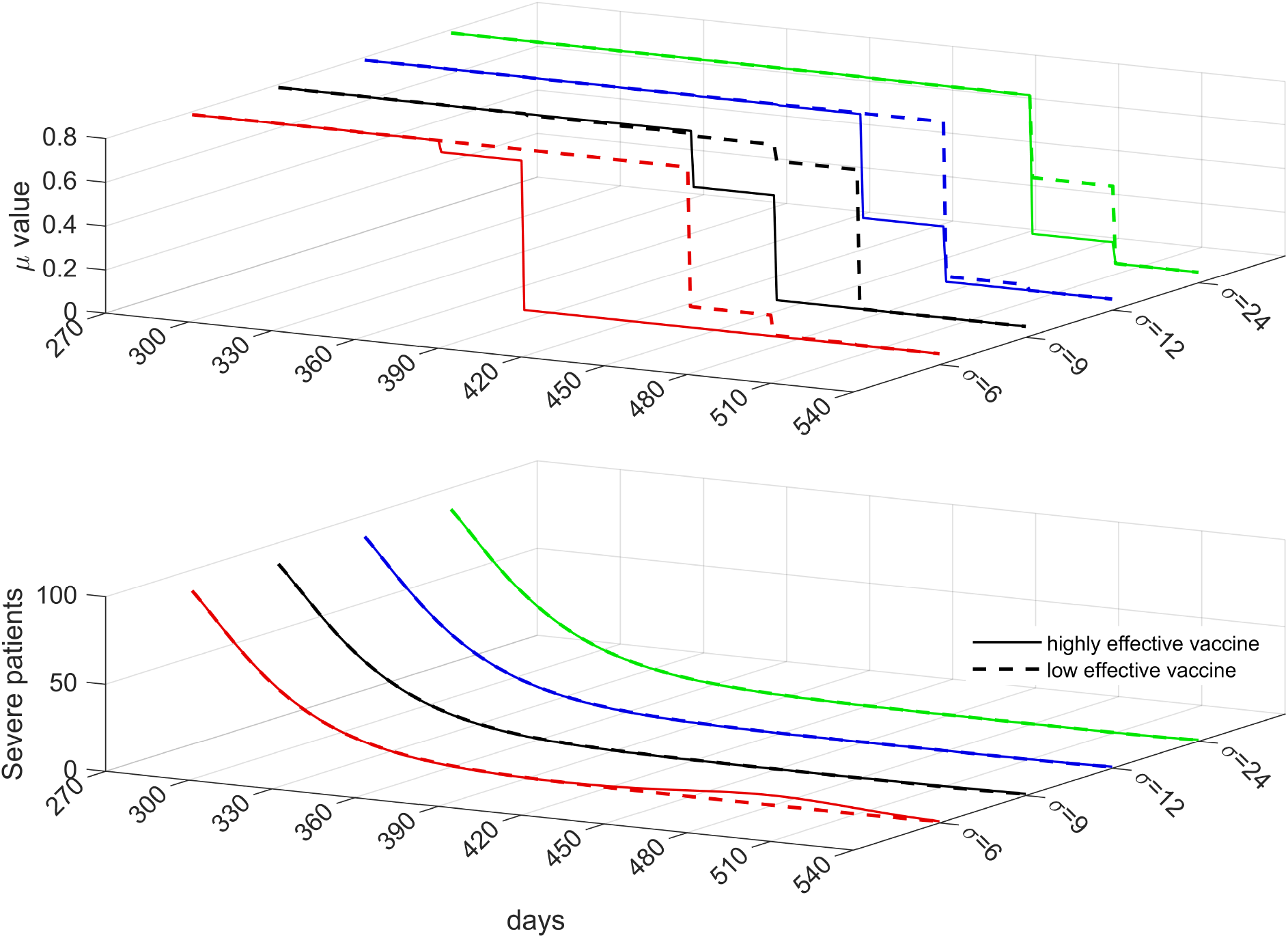
Optimal strategies considering different values for vaccine effectiveness and speed of vaccination. The optimal values of *µ* per month for highly (solid) and low (dashed) effective vaccines are shown on the top panel, while the corresponding number of severe patients is shown on the bottom panel. The red, black, blue, and green curves correspond to vaccinating 80% of the initial total population in 6, 9, 12, and 24 months, respectively.

If vaccination proceeds at a speed equivalent to vaccinating 80% of the initial total population in six months (red solid) using a highly effective vaccine, the optimal strategy is to keep the value of *µ* at around 0.785 from the start of the vaccination phase until day 359, when the first small noticeable reduction in *µ* to 0.733 is observed. This value of *µ* is reduced further to 0.05 on day 390. If the speed of vaccination is slowed down and vaccinating 80% of the initial total population is achieved in 9 (black solid), 12 (blue solid), or 24 months (green solid), the first observable easing of NPIs is delayed to day 419, 449, and 479, respectively. Despite the delay, the corresponding reduction in the value of *µ* is greater. The value of µ decreased to 0.530 (black solid), 0.303 (blue solid), or 0.146 (green solid). In all scenarios, the number of severe patients by day 540 is around 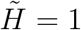, the threshold value set for the payoff constraint during the vaccination phase. Notably, a small peak in severe patients is observed on day 480 when the speed of vaccination is the fastest (red solid).

When a low effective vaccine is used, the first significant reductions in NPIs are further delayed compared to when highly effective vaccines were used. From around *µ* = 0.785 at the start of the vaccination phase, the value of µ noticeably reduced to 0.145 (red dashed) or 0.687 (black dashed) on day 450. Meanwhile, if the speed of vaccination is slower, the first noticeable reduction in µ is on day 480 to 0.075 (blue dashed) or 0.403 (green dashed). In all cases, the number of severe patients declined.

### 3.3. Effects of importation and initial number of infection on NPIs

Now we investigate the effects of importation and the initial number of infections on the optimization problem during the prevaccination phase. For simplicity, we assume that there is a uniform policy (*n*_1_ = 1) throughout the period. We vary the severe bed capacity *H*_*max*_, daily imported cases ξ, and initial number of infectious individuals *I*_0_.

On the left panel in Figure 7, we set *H*_*max*_ to 50, 100, 150, and 200, and vary the initial number of infected individuals *I*_0_. We assume that ξ = 0 and *E*_0_ = 10*I*_0_. The plot shows that for *H*_*max*_ = 150 or 200, the optimal values of µ range from 0.61 to 0.69 as the initial number of infections varies from 0 to 500. If *H*_*max*_ = 100, the optimal values of *µ* can climb to 0.7 or higher if I_0_ is at least 350. If *H*_*max*_ = 50 and the initial number of infections is at least 420, the maximum value for *µ* is reached.

**Figure 7:**
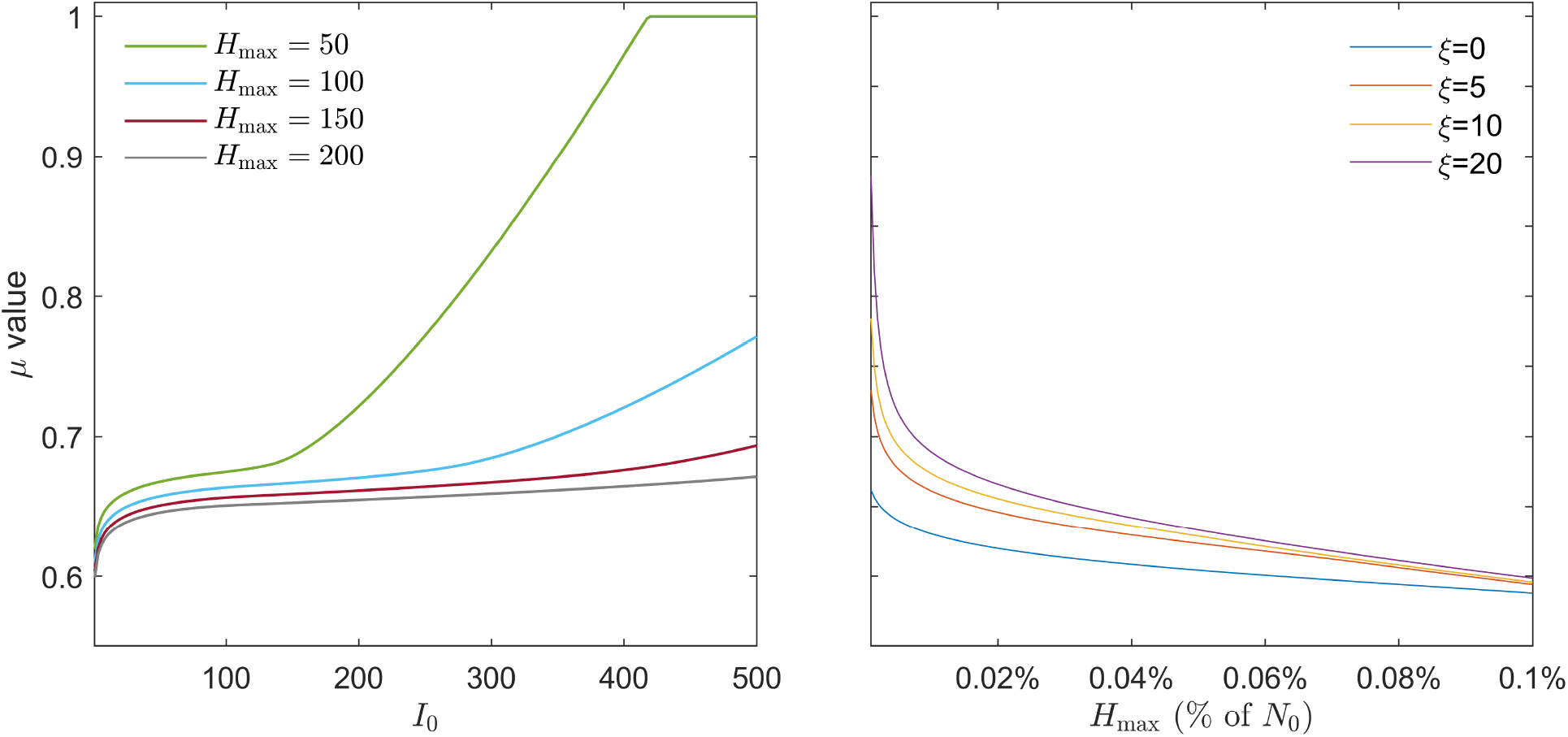
Optimal values of *µ* considering imported cases, initial number of infections, and severe bed capacity. The left panel shows the optimal value of *µ* for varying initial number of infectious individuals *I*_0_ and *H*_max_, while the right panel shows the optimal value of *µ* for varying severe bed capacity *H*_max_ and daily imported cases *ξ*.

On the right panel in Figure 7, we fix the daily number of imported cases ξ to 0, 5, 10, or 20, and vary the severe bed capacity *H*_*max*_. If *H*_*max*_ is only 0.001% of the total population, then the optimal values of *µ* are 0.818, 0.750, 0.714, and 0.657 if ξ = 20, 10, 5, or 0, respectively. Meanwhile, if the severe bed capacity is increased to 0.025% of the total population, then the optimal values of *µ* range from 0.689 to 0.704 as ξ varies from 0 to 20. As the capacity *H*_*max*_ is increased to 0.1% of the total population, all four cases converge to an optimal *µ* value of about 0.6.

### 3.4. Application of the framework using COVID-19 data of Korea

The estimation results for the values *µ* from February 26, 2021, when vaccination was begun, until February 3, 2022, when the testing method changed are shown in Figure 8 and summarized in Table 3. The lowest *µ* value was 0.590 during the GR phase, and the highest value was 0.795 during SD4. When we consider the average values in each phase, GR (0.623) was the most relaxed, followed by SD2 (0.665), SGR (0.682), and SD4 (0.744). If we compare the length of time for the variants to reach 90% of the infections, delta took 16 weeks while omicron took 10 weeks. At the final two weeks of the estimation period (January 19 to February 3, 2022), daily cases exceeded 30000 and *µ* was estimated at 0.653.

**Figure 8:**
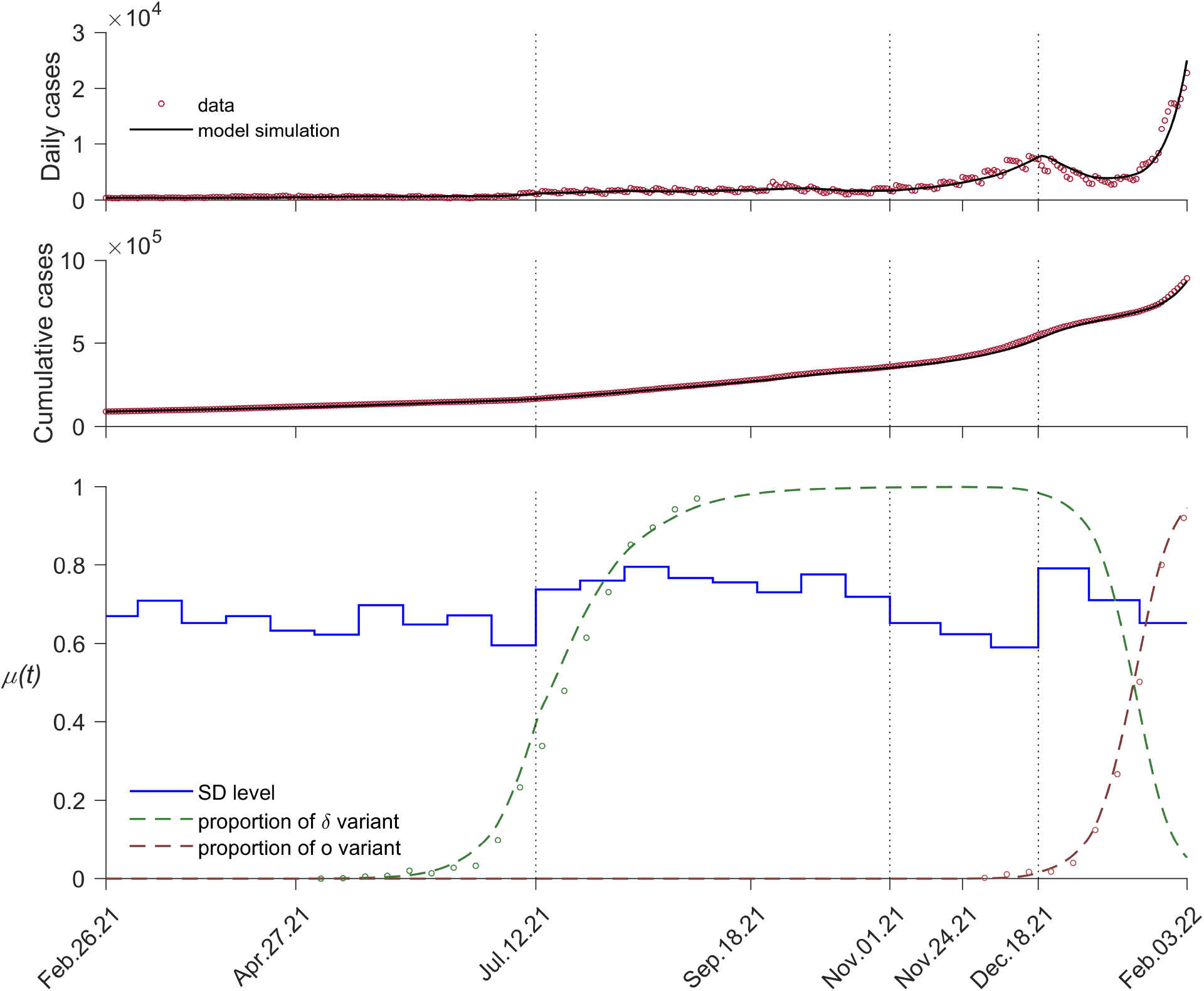
Fitting results. Model simulation (black curve) and data points (circles) for the daily (top panel) and cumulative (middle panel) cases. Bottom panel shows the fitted values of *µ*(*t*) (blue lines) and proportion of delta (green) and omicron (brown) variants.

**Table 3:** Estimated *µ*(*t*) values on each SD phase from February 26, 2021 to Februrary 3, 2022.

| Period | SD Policy | Average $\mu(t)$ | Range of $\mu(t)$ |
| --- | --- | --- | --- |
| Feb 26 to Jul 11, 2021 | SD2 (Level 2) | 0.665 | (0.596,0.737) |
| Jul 12 to Oct 31, 2021 | SD4 (Level 4) | 0.744 | (0.653,0.795) |
| Nov 1 to Dec 18, 2021 | GR (Gradual Recovery) | 0.623 | (0.590,0.653) |
| Dec 19, 2021 to Feb 3, 2022 | SGR (Suspended GR) | 0.682 | (0.653,0.791) |

Figure 9 shows the forecasts for the daily confirmed cases and severe patients under different levels of *µ* and amounts of antiviral drugs from February 3 to December 31, 2022. For this simulation, the amount of antiviral drugs is set to seven million (solid) or five million (dashed), and *µ* is kept constant until the end of the year. The red dotted line on the right panel shows the severe bed capacity at that time equal to 2825. Note that the drugs only affect the number of severe patients and not the daily confirmed cases since the drugs are assumed to reduce the severity of infections. If *µ* is fixed at 0.10 or 0.30, the number of severe patients surpasses the threshold. If the amount of antiviral drugs is limited to five million, a surge in severe infections starting from around July 2022 may occur if *µ* = 0.10, 0.30, or 0.50. Moreover, if *µ* = 0.10 and with five million antiviral drugs, the number of severe patients may once more exceed the threshold. In the next simulations, we apply the framework and investigate when and by how much should NPIs be eased given different amounts of antiviral drugs.

**Figure 9:**
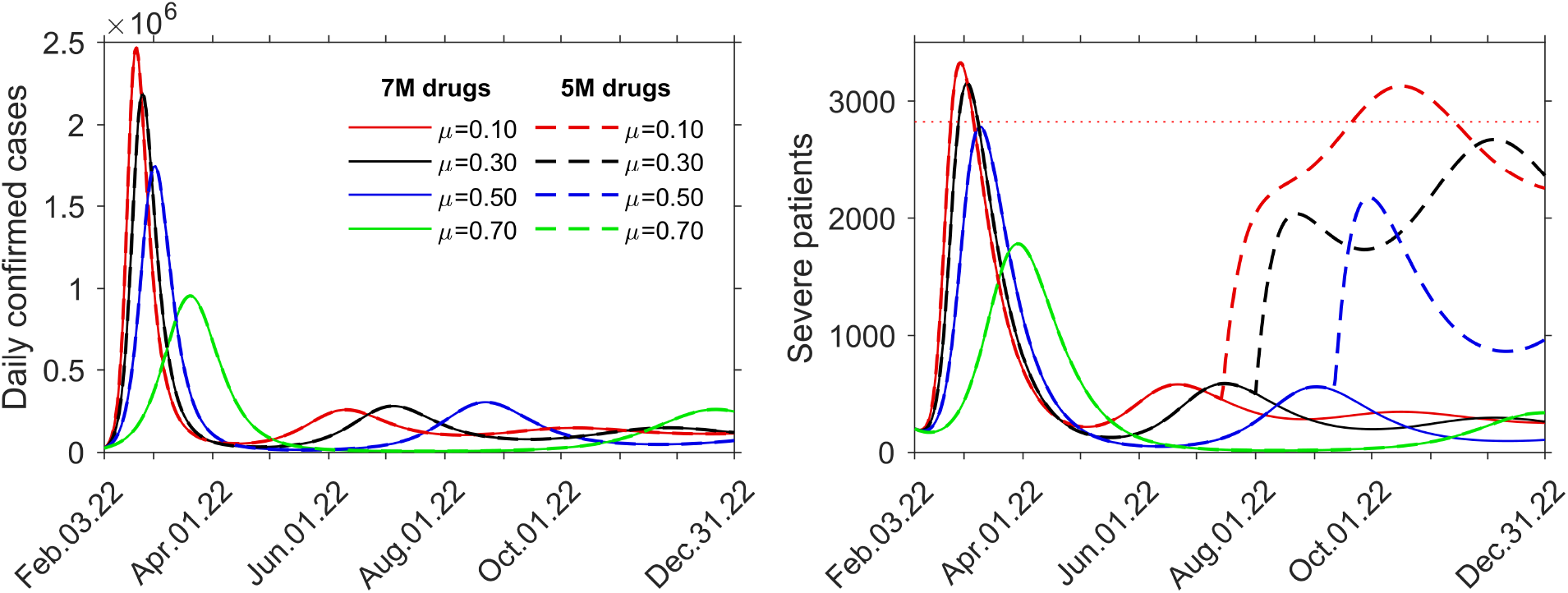
Daily cases and severe patients for different amounts of antiviral drugs and levels of NPIs. Red, black, blue, and green curves correspond to the constant values *µ* = 0.1, *µ* = 0.3, *µ* = 0.5, and *µ* = 0.7, respectively. The supply of antiviral drugs is set to seven million (solid) or five million (dashed).

Because Korea is currently in the vaccination phase, the optimal *µ* is solved by minimizing (11), where the number of severe patients is obtained from the solution of system (A.1). We set the upper bound for *µ* to 0.653, which was the last estimated value of *µ*, and set different amounts of antiviral drugs (five, six, or seven million). If the amount of drugs is five million (red), the optimal values of *µ* are around 0.6 until July 2022, then around 0.48 until the end of the year. A rise in severe patients may occur towards the last quarter of the year when the supplies are all used up. Meanwhile, if the amount of drugs is six million (blue), the optimal values of *µ* range from 0.487 to 0.438 until around October 2022 before it is reduced to 0.365. If the supply of antiviral drugs is seven million (green), the optimal values of *µ* are 0.534 at the beginning, eased to 0.318 from March 2022, then reduced significantly to 0.05 from April 2022. At the end of the forecast period, the cumulative severe cases are 18494, 11152, and 13233 for the five, six, and seven million supply of antiviral drugs, respectively. The corresponding CER values are 0.652, 0.493, and 0.124.

Figure 11 shows the optimal policy plans and the resulting number of severe and cumulative severe patients from February 3 until December 31, 2022, assuming different proportions *ϕ* of administered antiviral drug Paxlovid. Since the effectiveness of Paxlovid (89%) is assumed to be much higher than Lagevrio (30%), the net effectiveness of using both antiviral drugs would vary depending on how much of each drug is administered. For example, *ϕ* = 0.8 means out of all the administered antiviral drugs, 80% is Paxlovid and 20% is Lagevrio. This results in net effectiveness of 77%. In the forecast, the optimal values of *µ* on the first period if *ϕ* = 1 (green) or *ϕ* = 0.96 (teal) are 0.534 or 0.617, respectively. On the other hand, if *ϕ* = 0.92, 0.88, 0.84 and 0.8 the optimal values of *µ* are 0.674, 0.706, 0.728, and 0.743, respectively, which are higher than the estimated value of *µ* = 0.653 before February 3, 2022. At the end of the forecast period, the optimal value of *µ* reached the minimum (0.05) in all scenarios and the cumulative severe cases are 23861, 21815, 20100, 17922, 15656, and 13231 for *ϕ* = 0.8, 0.84, 0.88, 0.92, 0.96, and 1, respectively. The corresponding CER values are 0.290, 0.272, 213, 0.181, 0.166, and 0.124.

## 4. Discussion

In Figure 5, the oscillating values of *µ*, which results in the shifts in the number of severe patients, are observed on the quarterly, monthly, biweekly, and weekly policy changes. In the quarterly policy change (*n*_1_ = 3), relaxation of NPIs is suggested to be implemented from months 3 to 6. Correspondingly, we observe a very small number of severe cases until towards the end of the sixth month. Under this strategy, NPIs should be intensified in the last period as the number of severe cases increased and peaked at almost the capacity *H*_*max*_ before it declined. As the frequency of policy change is increased, there are more frequent adjustments in the intensity of NPIs, depicted by the jumps in the values of *µ*. Consequently, more peaks in the number of severe patients occur. Naturally, a rise or drop in cases follows when the policy is eased or relaxed. In all cases, *H*_*max*_ is always almost reached to allow the most relaxed level of NPIs possible, as long as the number of severe cases is kept below the capacity. Results of the cost-effectiveness analysis show that the monthly policy change has the least CER (0.012) and hence, the most cost-effective strategy. On the other hand, the weekly policy change is the least cost-effective (0.031).

In Figure 6, we see that a gradual easing of NPIs (or decreasing values of *µ*) is possible in all scenarios. We observe that NPIs can be noticeably eased much earlier if the vaccines used are highly effective. Moreover, if vaccination is slow (green curves), then strict NPIs (*µ ≈* 0.78) are maintained longer, even if the vaccine has high or low effectiveness (easing on day 479 for high and on day 480 for low vaccine effectiveness). These emphasize the importance of not only using highly effective vaccines but also fast administration of the vaccines to the population.

In Figure 7, results show that with higher *H*_*max*_, a lower value for the optimal µ is possible. For low values of initial infections (*I*_0_ less than 150), the level of NPIs is relatively stable (*µ* from 0.61 to 0.69). As I_0_ increases and *H*_*max*_ reduces, the importance of maintaining strict NPIs is more evident. For example, if *I*_0_ = 500 (or 0.05% of *N*_0_) and *H*_*max*_ = 50 (or 0.005% of *N*_0_), the healthcare capacity can be overwhelmed with severe cases even with the most strict level of NPIs (*µ ≈* 0.95). Thus, this approach can serve as a guide in assessing the adequacy of severe beds given that the initial number of infections is known. Furthermore, the impact of daily imported cases on the optimal values of *µ* is more apparent for smaller values of *H*_*max*_.

Here, the severe bed capacity *H*_*max*_ is presented as a percentage of the total population. For example, in Germany and the USA in 2020, the number of ICU beds is more than 0.025% of the population, while in most African and Southeast Asian countries, this percentage is less than 0.001% [50]. In Figure 7, if the severe bed capacity is 0.001% of the initial population, the intensity of NPIs is less if there are fewer imported cases (*µ* = 0.657 if ξ = 0, while *µ* = 0.818 if ξ = 20). Therefore, for countries with a low number of severe beds, NPIs such as screening measures at the border are crucial in keeping a minimal number of imported cases and preventing strict social distancing policies.

In the parameter estimation using COVID-19 data of Korea from February 26, 2021 to February 3, 2022, the estimated *µ* values and the number of daily confirmed cases during SD2 and SD4 were relatively stable except around July 12, 2021, when the proportion of the delta variant among the infections increased rapidly. On November 1, 2021, GR was implemented and the maximum number of people allowed in a private gathering increased to eight. As a result, daily confirmed cases soared, even though the proportion of omicron among the cases was still below 10%. As daily confirmed cases reached 7000, the government suspended the GR policy. Since SGR initially had a stricter private gathering policy, the daily confirmed cases decreased. However, the daily confirmed cases increased again as the proportion of the omicron variant rose to over 50%. Despite the increasing number of cases towards the end of the estimation period, policies in Korea kept easing.

Furthermore, we investigate the effects of antiviral drugs and the easing of NPIs in the number of severe patients. We see in Figure 9 that for non-optimal *µ* values of 0.1 or 0.3, even with seven million antiviral drugs, the number of severe patients may surpass the severe bed capacity. Moreover, with *µ* = 0.5 or lower and five million antiviral drugs, a surge in severe patients is likely to happen. Results in Figure 10 with the optimal µ values show that in all three cases, a gradual easing of NPIs is possible without exceeding the severe bed capacity, even during the second surge of cases when all the five million supply of antiviral drugs are used up. We note that all the obtained optimal *µ* values are less than the estimated *µ* values during SD2, SD4, GR, and GR (see Table 3). Although the cumulative cases by the end of the year when the antiviral drugs are six million (11152) is less compared to when the supply is seven million (13233), the CER for the seven million supply (0.124) are considerably less than the six million supply (0.493). Finally, we see on the bottom left panel in Figure 11 that if the proportion of administered Paxlovid is 92% or less (blue, black, purple, and red), then more strict NPIs, greater than the previously estimated *µ* = 0.653, should be implemented at the beginning of the forecast period so that the number of severe patients will not exceed the set threshold of 2825 beds. In all cases, a very relaxed level of NPIs (*µ* = 0.05) is possible starting June 2022. A lower proportion *ϕ* may delay the peak of severe infections but the wave is wider resulting to higher cumulative cases. For instance, if *ϕ* = 0.80, the forecasted cumulative severe cases is almost double compared to when *ϕ* = 1. This is interesting because social distancing policies are more relaxed if *ϕ* = 1 compared to when *ϕ* = 0.8. Thus, the use of highly effective antiviral drugs not only reduces the number of severe cases but may also lead to earlier easing and less strict intensity of NPIs. In Korea, COVID-19 restrictions were eased starting in April 2022, while the relaxation of mask mandates began in May 2022. Since 93% of antiviral drugs used in Korea were Paxlovid, the results of the simulations support the Korean policies on the easing of restrictions.

**Figure 10:**
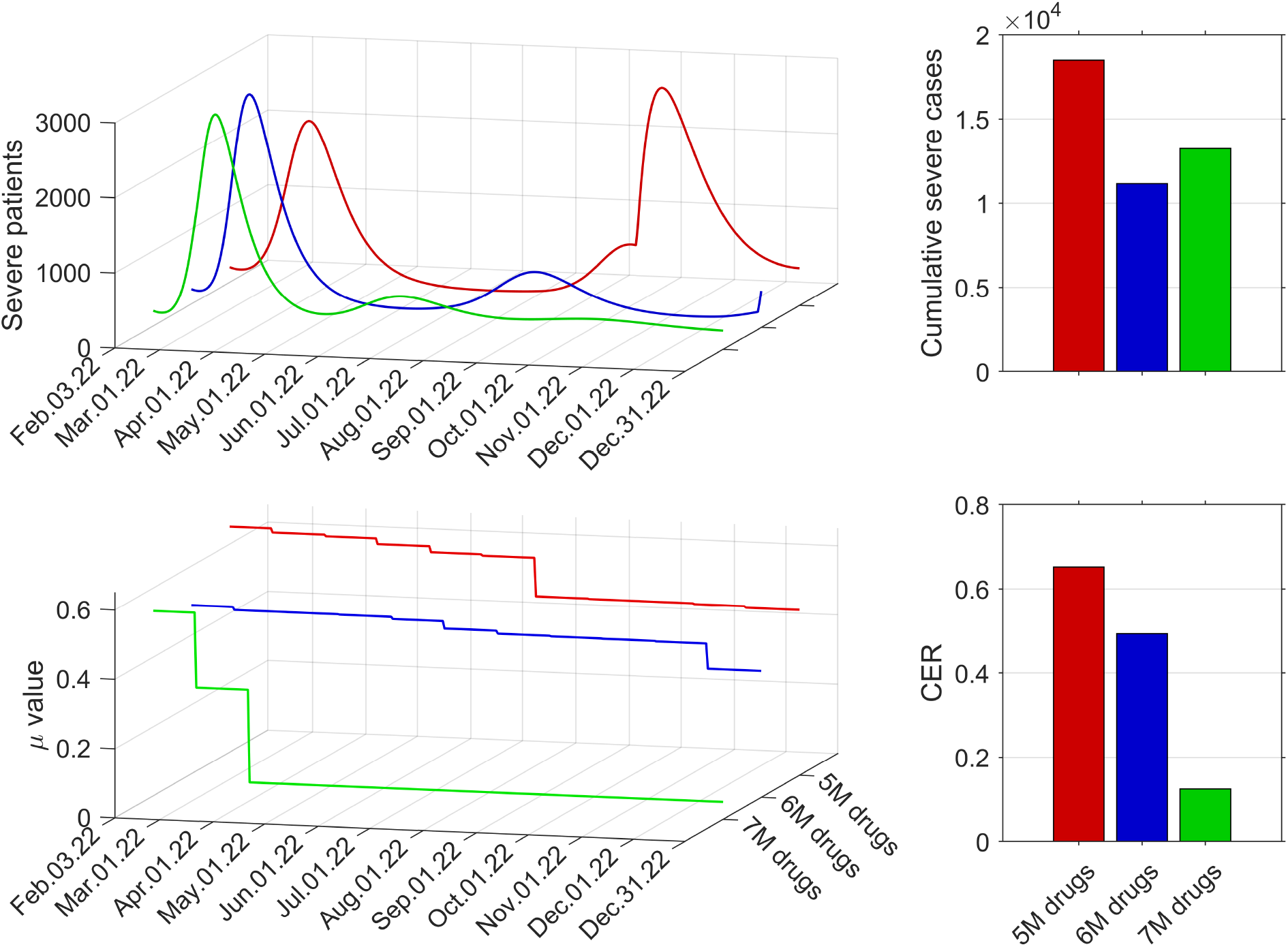
Forecast results using the optimal values of *µ* given different amounts of antiviral drugs. Panels on the left show the number of severe patients and optimal values of *µ* from February 3 to December 31, 2022, if the supply of antiviral drugs is five million (red), six million (blue), or seven million (green). The bar plots show the cumulative severe cases by the end of the forecast period and the corresponding cost-effectiveness ratio (CER) for five, six, or seven million antiviral drugs.

**Figure 11:**
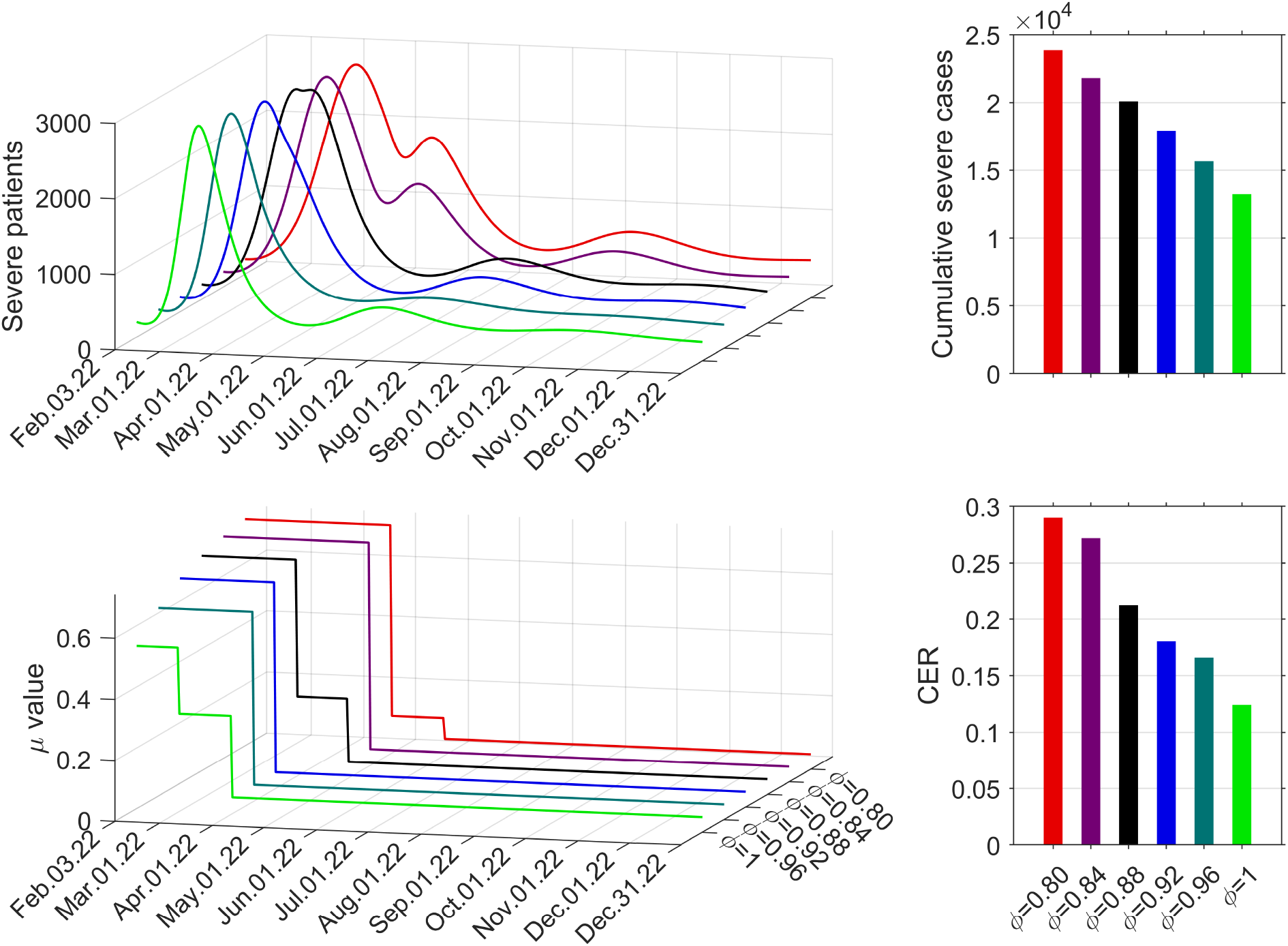
Forecast results using the optimal values of *µ* given different proportions of the antiviral drugs. Panels on the left show the number of severe patients and optimal values of *µ* from February 3 to December 31, 2022 if the proportion of Paxlovid *ϕ* used among the antiviral drugs is 80% (red), 84% (purple), 88% (black), 92% (blue), 96% (teal), or 100% (green). The bar plots show the cumulative severe cases by the end of the forecast period and the corresponding cost-effectiveness ratio (CER) for different values of *ϕ*.

## 5. Conclusion

In this work, we have formulated a nonlinear optimization problem that minimizes the intensity of NPIs (such as social distancing and mask-wearing) but ensures that the number of severe cases will not surpass the healthcare capacity. We consider two phases in the policy plan: prevaccination and vaccination. An epidemiological model that considers vaccination and quantifies the reduction in transmission due to NPIs (1*−µ*) and the number of severe cases 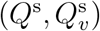 is embedded in the optimization problem. The severe bed capacity is incorporated as a constraint. The constrained optimization problem is transformed into an unconstrained one using the penalty method. Because we use exact penalty functions, the resulting objective function is non-differentiable, non-convex, and highly multi-modal. The metaheuristic optimization algorithm IMODE, which is capable of obtaining the global minimum, is used to solve the problem.

In the simulations, we set the length of the prevaccination and vaccination phases to 9 months. During the pre-vaccination phase, a payoff constraint is added that guarantees the decline of cases at the end of this phase. It is observed that more frequent policy changes result in more oscillations in the number of severe cases and a lower average *µ* on the entire phase. However, considering the cost of implementation of NPIs and the number of averted severe cases, results of the cost-effectiveness analysis show that a monthly policy change is the most cost-effective.

In the vaccination phase, we added constraints to ensure that gradual easing of NPIs as more people got vaccinated, and a target number of severe cases at the end of the vaccination phase is attained. Faster administration of vaccines results in the earlier easing of NPIs. If 80% of the population is vaccinated in 6 months, we have shown that NPIs can be eased 91 days earlier if highly effective vaccines were used compared to if low effective vaccines were used. Furthermore, we have seen that if vaccine rollout is slow (80% of the population is vaccinated in 24 months), vaccine effectiveness does not impact the timing of easing of NPIs. We also investigated the effects of importation and the initial number of infectious individuals on the optimal *µ* values. We have demonstrated that the initial number of infected individuals and daily imported cases should be kept at a minimum value especially when the severe bed capacity is low.

As an application, we used the optimization framework to determine optimal social distancing policy plans in Korea considering different amounts and types of antiviral drugs. A mathematical model that includes variants, booster vaccines, antiviral drugs, and waning of immunity is used. To establish the relationship between social distancing policies and the values of *µ*, the model is first fitted to the available data on confirmed cases by minimizing a least-squares formulation using IMODE. Forecast results for the optimal *µ* values show that gradual easing of policies is possible and the number of severe patients can be maintained below capacity even with at most 5 million antiviral drugs, compared to when *µ* is kept fixed to 0.3 or below. The easing of NPIs may occur earlier and with less intensity, if the supply of antiviral drugs is enough and the antiviral drugs are highly effective in reducing the severity of the disease.

The study of evolutionary algorithms is an active research area. Other recent algorithms can be explored to solve the optimization problem. For future work, one can do a comparative study on different optimization algorithms that can solve the minimization problem with the fastest time and highest accuracy. In our simulations, a constant value for the severe bed capacity *H*_*max*_ was assumed. However, the optimization problem can be reformulated to allow *H*_*max*_ to be time-dependent, as in the case when there is a surge of infections and hospital bed capacity needs to be increased to accommodate more patients. A study that considers a timedependent *H*_*max*_ can be done in future work. One can also consider a more general optimization problem that allows for a non-constant duration of policies (e.g. 𝒫_1_ is 30 days, 𝒫_2_ is 15 days, and so on). Furthermore, we assumed that both the prevaccination and vaccination phases span 9 months. Depending on the capability to implement and availability of resources of a region or country, the length of these phases can be modified, and consequently, the optimal strategies will change. In the application using COVID-19 data of Korea, second booster shots and possible underreporting of cases were not considered. Nevertheless, the epidemiological model can be modified to incorporate these and other factors, and the framework is still applicable. The proposed scheme is general enough that it can be applied to any model, regardless of complexity, as long as NPIs are quantified as a reduction in the force of infection, and the severe case has a separate compartment. Finally, the presented framework may be applied to other variants of COVID-19 or other infectious diseases.

## Data Availability

The dataset for the number of confirmed COVID-19 cases in Korea supporting the conclusions of this article is available in the Korea Disease Control and Prevention Agency repository, http://ncov.mohw.go.kr/tcmBoardList.do?brdId=3&brdGubun= .

## Declarations

### Ethical Approval and Consent to participate

Not applicable

### Consent for publication

Not applicable

## Authors’ contributions

VMPM, RM, and JL - conceptualization, draft preparation, methodology, and software. EJ - conceptualization, draft preparation, methodology, project supervision. All authors read and approved the final version of the manuscript.

## Acknowledgements

Not applicable

## Appendix A. Equations of the COVID-19 model in the application using data of Korea

Below is the system of equations describing the mathematical model of COVID-19 in the Republic of Korea. Table A.4 shows the model parameters and their values considering the pre-delta, delta, and omicron variants.

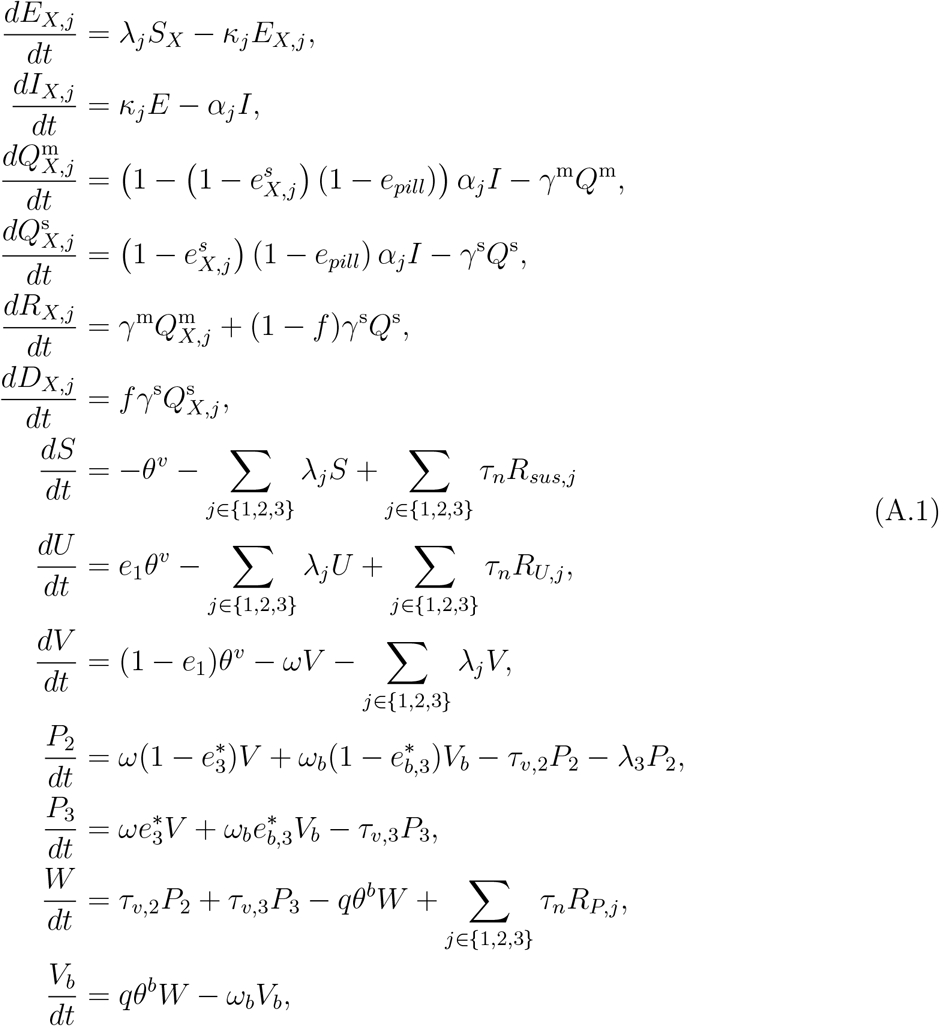

where

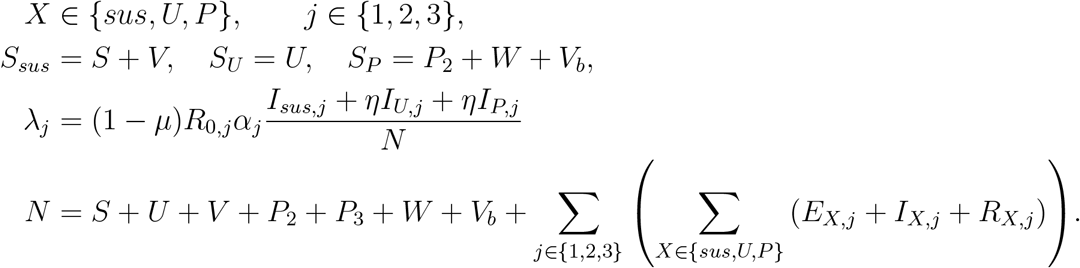

**Table A.4:**
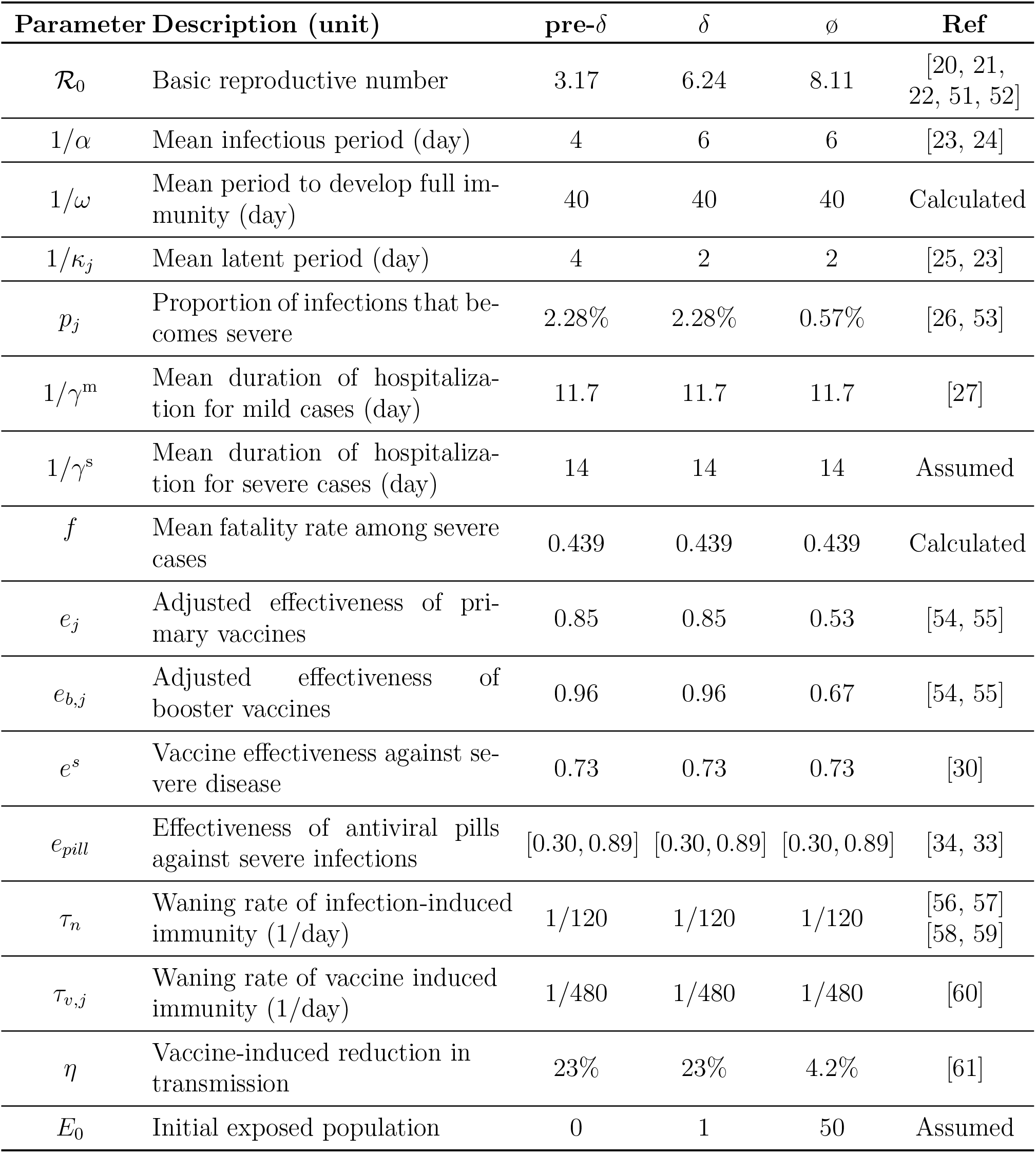
Description and values of the parameters in the mathematical model for application to Korea.

